# Initial Technical and Clinical Validation of Mobile Pupillometry with Virtual Reality: A Digital Biomarker for Screening Cognitive Function and Impairment

**DOI:** 10.64898/2026.07.15.26358187

**Authors:** Andy Brendler, Julia Fietz, Anastasia Bauer, Daniel Pfahl, Sarah Higgins, Eva Vidovic, Tanja Brückl, BeCOME Working Group, Memory Clinic Working, Kerstin Hupe, Matthias Knop, Victor I. Spoormaker

**Author notes:** Corresponding author: Andy Brendler, (or); Victor I. Spoormaker.

## Abstract

Cognitive impairment is a prevalent symptom extending from physiological ageing to disease. It commonly manifests itself in initial memory problems, progressing and co-occurring in more severe conditions such as Mild Cognitive Impairment, Alzheimer’s Disease and Major Depressive Disorder. However, current non-invasive screening assessments either lack biological information or are invasive and restricted to specialized centers with complex and cost-intensive set-ups. Here, we conducted an initial validation of mobile pupillometry with Virtual Reality (VR) under experimental conditions as a digital biomarker for cognitive impairment by testing required biomarker-specific properties. For this purpose, we first assessed its construct validity by testing healthy participants (n=43) on an n-back task in VR while pupil size was measured. Mixed effects models revealed that similar to lab-based eye-tracking systems, pupil size increased in a sensible and distinguishable fashion as a function of working memory load. Second, to test the signal’s reliability, the same participants were tested on the identical set-up two to three months after their first visit. We observed that the pupil response profile was highly stable over this period. Third, for its clinical validity, we examined patients (n=89) from three different cohorts with varying degrees of cognitive impairment and compared them to healthy control participants (n=81). Mixed-effects models indicated that pupil size was reduced as a function of cognitive impairment levels at higher cognitive load and that this effect was stronger pronounced with increasing age. In conclusion, we provide initial evidence for mobile pupillometry being a sensitive, reliable and clinically valid digital biomarker for cognitive functioning and impairment, which offers desirable properties due to its quick, automatized and location-independent set-up.

## Introduction

Healthy, intact cognitive functioning is a vital process for navigating through and managing our complex activities of daily living and has been associated with a number of positive outcomes including better health and greater well-being (John et al., 2025; Jokela et al., 2010). While cognitive functioning varies greatly among individuals, it is part of normative development that cognitive performance declines continuously over the course of life (Whitley et al., 2016). However, besides physiological decline in cognitive functioning, the transition to more pathological states is continuous and may develop gradually (Raimo et al., 2024). What often presents as an experience of subjective memory complaint may mark the onset of what later develops into more objective impairments, summarized under the umbrella term of cognitive impairment (Petersen et al., 2014; Rostamzadeh et al., 2022). Cognitive impairment – a transdiagnostic feature of Mild Cognitive Impairment (MCI), Alzheimer’s disease (AD) and Major Depressive Disorder (MDD) (Richard et al., 2013), among other neuropsychiatric conditions – is commonly defined as a reduction in mental processes such as memory, attention, and executive function, and is particularly evident in working memory deficits (Millan et al., 2012). However, as argued below, a highly accessible digital biomarker – defined as an objective and quantifiable physiological measurement acquired by digital devices (Babrak et al., 2019; Coravos et al., 2019; Vasudevan et al., 2022) – that is sensitive enough to capture the cognitive variability spectrum in pathological conditions such as MDD and AD has yet to be found. Thus, it requires a quantifiable digital biomarker, which allows to characterize the gradient from healthy cognitive functioning, through slowly deteriorating cognitive states starting with cognitive impairment and transitioning into or co-occurring with more severe pathological conditions.

This is especially important as MDD and AD are among the most pressing global health concerns with an estimated rise in cases in the upcoming years (GBD Collaborators Mental Disorders, 2022; Nichols et al., 2022) .These trends pointing to a rapidly rising number of cognitively impaired individuals inevitably will require new screening tools that can aid the healthcare system in managing an increasing demand in diagnostic workload. Even though current diagnostic biomarkers for Alzheimer’s disease – reduced cerebrospinal fluid (CSF) amyloid-beta 42 (Aβ42) concentrations, mostly determined by the Aß42/Aß40 ratio (Jack et al., 2018), evidence of tau pathology (Hansson, 2021) or increased cortical amyloid accumulation detected by positron emission tomography PET; Villain et al., 2012) – are diagnostically sensitive and also predictive of disease progression, their application is limited due to their invasiveness, high cost, and restricted accessibility to specialized clinical settings. This restricts the screening potential to a limited number of affected people for whom the disease has often already reached a clinically significant stage at the time of testing (Jia et al., 2024). Recent developments in the biomarker field come closer to address these disadvantages through blood-based biomarker (Palmqvist et al., 2024) that are much less expensive, time-consuming and more accessible to a larger number of healthcare providers. In particular, plasma phosphorylated tau 217 (p-tau217) showed desirable properties with strong association to CSF markers. However, as these tests are not intended to quantify cognitive status including cognitive impairment directly, additional biomarkers targeting this domain are needed. This is further strengthened by the fact that initial evidence (Palmqvist et al., 2024) indicated that these promising blood biomarkers have increased diagnostic accuracy if information on cognitive impairment status can be utilized. In contrast, for MDD reliable biomarkers that are part of common diagnostic practices remain to be determined.

One cognitive process that has been shown to be involved in cognitive impairment is working memory – the cognitive system that allows to hold information in mind for a short-time interval while simultaneously processing it (Baddeley, 2012). Physiologically, working memory relies on sustained arousal and neuromodulatory input, particularly from the noradrenergic and dopaminergic systems, to maintain and manipulate information over short periods during ongoing cognitive processing (Arnsten, 2009; Sara & Bouret, 2012). This arousal-linked cognitive control has been shown to be disrupted in both depression and early AD, suggesting dysfunction of the frontoparietal and fronto-striatal circuits that support working memory, and thus serving as a potential early indicator of these conditions (Petersen, 2004; Snyder, 2013). MCI, considered a prodromal phase of AD, is also frequently observed in late-life depression, highlighting the comorbidity of both disorders (Geda et al., 2006). Effective biomarkers for these conditions should be sensitive to early functional disruption of cognitive processes and specific to underlying neural systems (Hampel et al., 2018). However, many current digital markers (e.g., smartphone apps, wearables) fall short (Coravos et al., 2019), as they typically lack access to higher-order cognitive systems or fail to capture dynamic neurophysiological processes such as arousal-linked modulation of working memory performance.

Noteworthy, in this context, a converging brain structure that has been observed to be altered in AD and MDD and being linked to cognitive functioning (Negelspach et al., 2025) and arousal regulation, is the locus coeruleus (LC) – a nucleus located deep within the brainstem that is rich in noradrenergic neurons and contributes to widespread noradrenaline (NA) signaling across higher-order brain regions (Poe et al., 2020). The LC is among the first structures being affected of elevated tau protein accumulation in early stages of AD (Gutiérrez et al., 2022), and also in stress-related mental disorders such as MDD, altered LC activation has been observed and linked to aberrant brain network dynamics (Del Cerro et al., 2020; Maturana-Quijada et al., 2024; Slavova et al., 2024).

One physiological measure indirectly capturing LC activity dynamics constitutes experimentally-probed brain arousal assessed with pupillometry – the method of measuring pupil size changes during higher-order processing contexts (Strauch et al., 2022). Here, it is of crucial importance to emphasize task engagement as pupil size fluctuations can also be assessed during rest, presumably tapping into other LC regulated activity patterns (for an elaborate discussion of LC’s activity profile regulation, see Aston-Jones & Cohen, 2005; Joshi et al., 2016; Murphy et al., 2014; Strauch et al., 2022).

A series of laboratory studies including simultaneous fMRI/pupillometry studies could show that experimentally probed brain arousal originating from the LC and modulated by higher-order areas such as the salience network (Lee et al., 2020; Ross & Van Bockstaele, 2021) can be assessed by pupillometry measurements (de Gee et al., 2017; Fietz et al., 2022; Murphy et al., 2014; Weiss et al., 2026) with these findings showing reproducible sensitivity to changes across healthy and clinical populations. In this context, a consistent finding constitutes that pupil size and dilation as surrogate marker of brain arousal levels – most probable mediated through LC activity regulation processes – is reduced across reward and cognitive domains in subgroups of neuropsychiatric patient cohorts (Brendler et al., 2024; Fietz et al., 2024).

These consistent findings make experimental pupillometry a promising candidate read-out to serve as a biomarker for cognitive impairment. However, current findings in this context are almost exclusively based on rigid laboratory-based set-ups in which pupillometry data is acquired in a highly controlled but static environment. As outlined before, the high number in anticipated neuropsychiatric patient cases requires a biomarker that is non-invasive, quick in testing and aligns with existing neurobiological findings in MCI, AD and MDD. In addition, and most importantly to meet the high demand of diagnostic testing required, it is a mobile biomarker solution that would allow to increase the process and frequency of early testing in suspected cases. However, to date, such a mobile biomarker for quick diagnostics **–** one that would open the avenue for widespread and location-independent testing while offering meaningful physiological information on the early progression of cognitive impairment **–** has not been validated.

In order to address this gap, with the goal to further characterize and validate mobile pupillometry with VR as a digital biomarker for cognitive impairment, we assessed three fundamental biomarker-required properties of this measure: First, in a group of healthy participants (HC), we studied the construct validity by testing if our mobile measure yields the expected scaling of pupil size with increasing cognitive load. Furthermore, to test the stability of this measure over time, we assessed its test-retest reliability re-assessing the same group of healthy participants after a time interval of two to three months. Finally, to examine how well it performs in terms of its clinical validity, we explored patients with varying cognitive impairment levels along a cognitive functioning gradient ranging from relatively unimpaired to strongly impaired and compared their pupillary response profiles to healthy control participants.

## Materials and methods

### Participants

For investigating the construct validity and intra-participant reliability of arousal-assessed cognition with mobile pupillometry, we analyzed data from 43 healthy participants (age range: 21 **–** 65 years, age *M* = 31.6, *SD* = 11.3, 27 women), who were invited to the psychophysiology laboratory at the Max Planck Institute of Psychiatry in Munich. After 60 to 90 days following their first visit, all participants were invited for a second visit to complete the exact same experimental procedure. In total 33 participants (age range: 21 **–** 65 years, age *M* = 31.9, *SD* = 11.7, 21 women) took part in this follow up assessment (**Figure 1b**).

**Figure 1.**
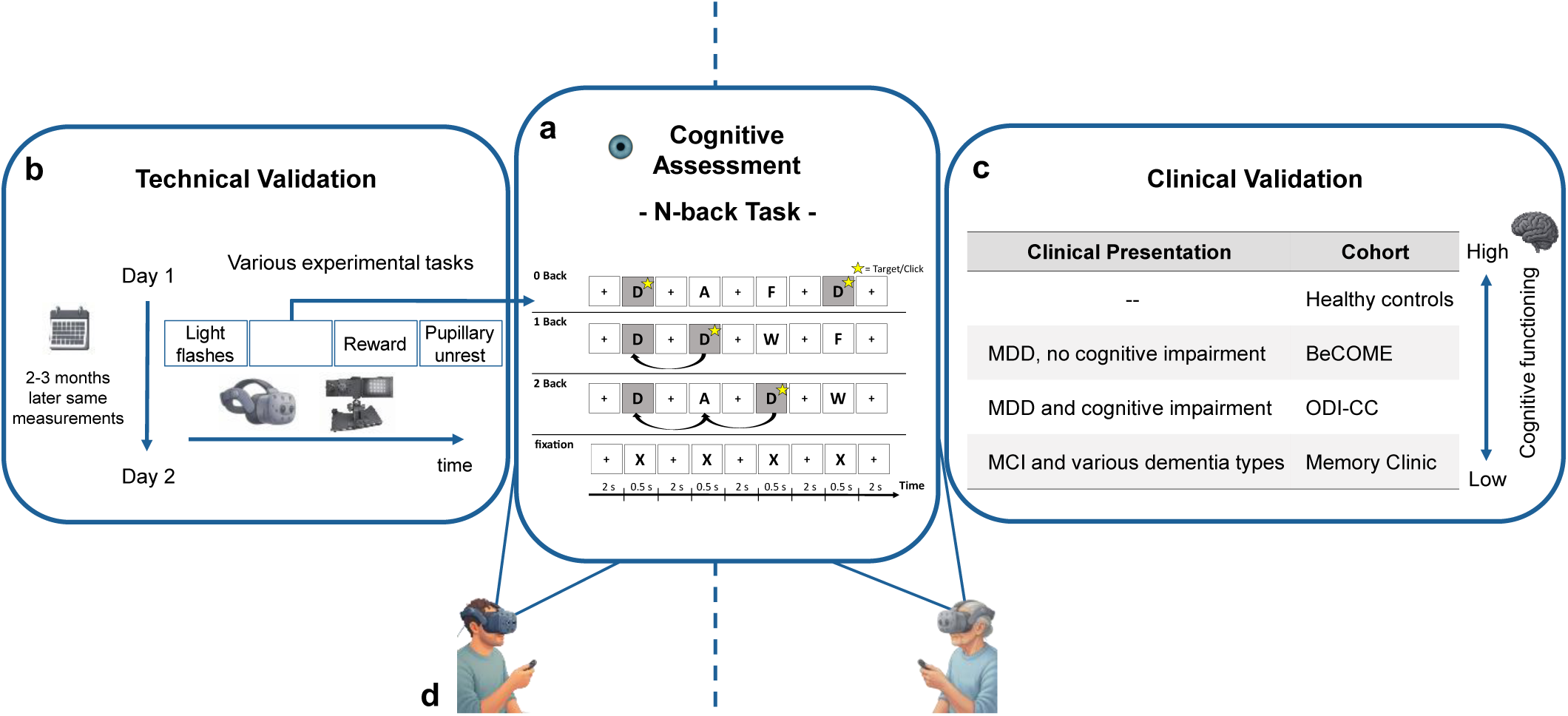
Schematic depiction of the experimental procedure. **a.** A visual depiction of the n-back task, the main task of this study **b.** Participants performed a series of experiments in VR: light flash, n-back, reward task, pupillary unrest and followed the exact same procedure 60 to 90 days after the first visit. **c.** Patients from different cohorts and healthy controls performed the n-back in VR **d.** Healthy participants and patients sat on a chair while wearing the VR headset and pressing a button.

Given the promising findings of this technical validation of our digital biomarker (see results), we aimed at evaluating its clinical validity by subsequently recruiting and testing patients from three distinct clinical cohorts using the same VR set-up (see **Table 1** for demographic and clinical details). The first group of patients (n=23) were inpatients on a ward specializing on affective disorders in the elderly (aged 60 or above), particularly with neurological comorbidities such as mild cognitive impairment. This cohort of older adults with depression and cognitive complaints, such as memory difficulties, who were receiving inpatient care is abbreviated as ODI-CC throughout the following sections. The second group of patients (n=53) was selected from the Biological Classification of Mental Disorder (BeCOME) study (Brückl et al., 2020) in which the VR measurements (described below) have been introduced in January 2025 after this earlier successful technical validation. Assessing patients from the BeCOME cohort allowed us to include a patient group characterized by mild affective symptoms with a relatively high level of functioning as these patients manage their daily lives independently and are mostly treated on an outpatient basis. The third and most severely cognitively affected patient group was recruited from the MPIP Memory Clinic (n=13), which focuses on memory complaints and neurodegeneration in early stages with suspected and/or confirmed dementia cases including AD (n=4). In addition, in total, we recruited 81 healthy control participants (of which 43 were identical with those reported for the technical validation and 16 participants aged 58 years or older were included as healthy elderly controls). These different patient cohorts, together with healthy control participants, allowed us to evaluate mobile pupillometry as a digital biomarker across a spectrum of cognitive impairment. The sample ranged from individuals with normative cognitive function, to high-functioning affective disorders patients from the BeCOME study, to more severely cognitively affected ODI-CC and Memory Clinic patients, including some diagnosed with dementia such as Alzheimer’s disease (**Figure 1c**). This gradient of cognitive impairment provided a well-suited framework to assess the clinical validity of this potential digital biomarker.

**Table 1.** Clinical and demographic characteristics of cohorts.

| Characteristic | HealthyControls<br>(n=81) | BeCOME<br>(n=53) | ODI-CC<br>(n=23) | Memory Clinic<br>(n=13) |
| --- | --- | --- | --- | --- |
| Age, M (SD) | 37.1 (15.7) | 35.8 (13.7) | 65.9 (7.8) | 74.1 (8.6) |
| Sex, n |  |  |  |  |
| Female | 50 | 40 | 10 | 5 |
| Male | 31 | 13 | 13 | 7 |
| Clinical Diagnosis, n |  |  |  |  |
| Major Depressive Disorder (MDD) | - | 42 | 21 | 5 |
| SCD | - | - | 1 | 1 |
| MCI | - | - | - | 7 |
| Dementia | - | - | 1 | 5 |
| Other mental disorders | - | 63 | - | - |

*Note.* Clinical and demographic characteristics of cohorts. Patients may have multiple diagnosis, thus, categories are not mutually exclusive. Dementia: Alzheimer's Disease, Vascular dementia; Other mental disorders: Dysthymia, Somatization disorder, Psychotic spectrum, Substance Use Disorders, Anxiety disorders, Post-Traumatic Stress Disorder.

The study procedure adhered to the principles of the Declaration of Helsinki and received approval from the local ethics committee at Ludwig Maximilian University of Munich. All participants were reimbursed either monetarily or with a voucher after their study participation. For all healthy participants, eligibility criteria included proficiency in either English or German, the absence of any prior diagnosis of neurological, neurodegenerative, or neuropsychiatric conditions, any former history of ocular disease and no current use of psychotropic medications. For patients, the criteria were identical expect that we specifically recruited participants with a suspected or confirmed diagnosis of depressive disorder, mild cognitive impairment, or Alzheimer’s disease. Furthermore, as one of the tasks included bright flashes of light, we specifically ensured that participants did not have a history of conditions sensitive to these stimuli (e.g., migraine or epilepsy).

### N-back task

In the present study, we used the n-back task (**Figure 1a**) – a well-established experimental task for probing working memory (Owen et al., 2005) – and operationalized cognitive load by presenting four different conditions with increasing difficulty respectively cognitive load levels (adapted from: Brückl et al., 2020; Fietz et al., 2022). During the fixation condition, which was the task baseline, participants had to only focus on the letter X presented on the screen without any required response or working memory component. In contrast, in the 0-back condition, participants had to press a button when a pre-specified target letter appeared on screen. In the 1-back condition, participants had to press the button as fast as possible when the letter on screen was the same as the letter presented before. In the 2-back condition, which was the one with the highest cognitive load, participants had to press the button when the letter on screen was the same as the one preceding the previous letter (i.e., two letters before the currently presented one). Each condition block lasted approximately 40 seconds and included an alternating presentation sequence of one letter at a time for 2 seconds, followed by a fixation cross for 0.5 seconds. This yielded a total of 16 letters per block with 4 targets (i.e., letters that required a response as fast as possible) and 8 non-targets (i.e., no response was required). All four condition blocks were presented twice, yielding a total of eight condition blocks per experimental session. For testing the clinical validity, we added one additional 1-back block in the beginning that served as practice block.

### Procedure

Participants were comfortably seated in a quiet room during the entire experiment. Pupillometry data were acquired using an HTC VIVE Pro Eye VR headset, which is equipped with an integrated Tobii eye-tracker capable of capturing high-resolution gaze and pupil diameter data with a sampling frequency of 120 Hz. The headset was securely and comfortably fitted for each participant, with adjustments made to ensure optimal eye-tracking calibration. Furthermore, participants received a pointer with which button presses for the experimental task were recorded (**Figure 1d**). Calibration was conducted at the beginning of the session using a standard five-point calibration protocol. Following calibration, participants completed three tasks presented in a fixed order: first, a series of five brief light flashes were presented to assess basic pupillary light reflexes. This was followed by the n-back task, the main experiment of the present study (**Figure 1a**). Lastly, the participants performed a reward task (for a detailed description of the task, see Brendler et al., 2024) followed by a resting period. All experimental tasks were presented on a virtual board within a straightforward direction relative to the participants first-viewer perspective; thus, no external head or body movement in this relatively static setting was required. This VR environment to present the experimental tasks was developed in an extension of previous work (Binder & Spoormaker, 2020).

After completing the VR-based assessments, some participants took part in an additional study set-up during which they were seated in front of a desktop-mounted eye-tracker while the pupil signal was recorded. This additional study set-up is not part of the present study. The exact study procedure was repeated with the same participants 60 to 90 days after their first visit (**Figure 1b**). For all patients from varying cognitive functioning cohorts and healthy controls, the VR procedure including the main n-back task was conducted (**Figure 1c**). Across all sessions and conditions, none of the participants reported motion sickness, discomfort, or other adverse effects associated with the use of the VR headset or the experimental procedures.

### Data pre-processing and statistical analysis

For data wrangling and pre-processing of the pupil and behavioral data, we used Python (version 3.14, Python Software Foundation) and Matlab (MathWorks, Natick, USA). For each participant, missing values in the pupil signal due to blinks were linearly interpolated using valid samples within a 100 ms sliding window preceding and following the missing data point. Raw pupil size values outside of a plausible biologically range (i.e., 2 mm **<** pupil value < 8 mm) (Mathot, 2018) were discarded and also interpolated. In order to account for inter-individual baseline differences in pupil size, the time series for each participant was standardized using z-transformation. Lastly, all data were also visually inspected to ensure high pupil data quality. We calculated the mean pupil size over time for each block within the different block conditions. Then, we averaged the values for blocks of the same condition, resulting in a single mean pupil size score for each of the four cognitive load levels. Similarly, for the behavioral data, we computed reaction time (RT) means for the target stimuli for each condition block. Accuracy was defined as the ratio of the total number of hits (i.e., correctly pressing a button in response to a target) to the total number of target stimuli presented.

For statistical analysis, we used the statistical programming language R (R Core Team, 2020). Given the repeated structure of our data, i.e., each participant contributed four observations for pupil data and three for the behavioral data (that is, four cognitive load conditions: fixation, 0-back, 1-back and 2-back, where fixation was omitted in the behavioral analysis as no response was required), the assumption of independence was violated. Hence, we fitted several linear mixed-effects models (LMMs) using the lme4 R package (Bates et al., 2015). In order to assess the construct validity of our measure, we fitted two LMMs, one with pupil and another with reaction time data as dependent variable. Furthermore, we fitted a generalized linear mixed-effects model (GLMM) for the accuracy data as dependent variable. For all models, restricted maximum likelihood estimation (REML) was used with a model structure, which was the same for all three models by specifying fixed effects for fixation, 0- back, 1-back and 2-back conditions, where the fixation condition was omitted in the behavioral models. Furthermore, for all models, a random intercept structure was used and the contrasts were set to sum-to-zero. Significance of fixed effects was evaluated using Type III F-tests with the Kenward-Roger approximation for degrees of freedom as implemented in the lmer package. Post-hoc contrasts were performed using the emmeans package (Lenth, 2021). The GLMM for the accuracy data was fitted using a binominal distribution and Laplace approximation using the Nelder-Mead optimizer. Post-hoc comparisons between conditions were done on the log-odds scale with Tukey adjustment for multiple comparisons. The model output is provided using odds ratios (OR), z-values and p-values.

To assess the test-retest reliability of mobile pupillometry, we calculated a differential score as the difference between the 2-back and 0-back conditions and computed the Pearson correlation coefficient between day one and day two measurements (i.e., 60 - 90 days apart). Furthermore, to assess the ranking of pupil scores within participant group over the two assessment days, we determined the Spearman’s Rank Correlation Coefficient. Finally, we computed the Intraclass Correlation Coefficient (ICC) based on a two-way mixed-effects model with absolute agreement (ICC(3,1)). The ICC is an index for assessing the degree to which measurements under similar conditions are consistent. This metric provides values between 0 and 1, with 1 indicating perfect reliability. There are different guidelines, which provide slightly different ICC interpretation reference values. According to Koo and Li (2016), values less than 0.5 indicate poor reliability, values between 0.5 and 0.75 indicate moderate reliability, values between 0.75 and 0.9 indicate good reliability, and values greater than 0.9 indicate excellent reliability. Cicchetti (1994) proposed the following cut-offs: values below 0.4 indicate poor, values between 0.4 and 0.59 fair, values between 0.6 and 0.74 good, and values 0.75 or above excellent reliability.

We examined the clinical validity by fitting two LMM’s using REML with the lme4 package. The first model included fixed effects for cognitive load (fixation, 0-back, 1-back, 2- back), group (HC, BeCOME, ODI-CC, Memory Clinic), and their interaction, with random intercepts for participants. We set HC as the reference level in order to allow comparisons to be interpreted relative to healthy cognitive functioning. As we were interested in the influence of age on pupil responses and groups, the second model had the exact same structure except that we included age after centering and modelled the three-way interaction of condition × group × age with a random intercept structure. Significance of fixed effects was evaluated using Type III F-tests with Kenward-Roger approximation for degrees of freedom to evaluate statistical significance.

## Results

### Construct validity of cognition-assessed mobile pupillometry

In order to test the construct validity of mobile pupillometry with VR, i.e., to which extent pupil size scales with cognitive load as commonly found in lab-based eye tracking systems, we fitted a linear mixed effect model for pupil size (**Figure 2a**). Pupil size increased significantly as a function of cognitive load (*ß =* 0.43 *SE* **=** 0.02*, F*(1, 131) = 543.03, *p <* .001) with each cognitive load level (*M*_fixation_ = -0.405, *SE*_fixation_ = 0.04, *M*_0-back_ = -0.040, SE_0-back_ = 0.04, *M*_1-back_ = 0.291, *SE*_1-back_ = 0.04, *M*_2-back_ = 0.923, *SE*_2-back_ = 0.04) showing significantly increased pupil size to the previous level (*ß*_0-back – fixation_ = 0.365, *SE* = 0.056, *t*(129) = 6.46, *p* < .001, *ß*_1-back – 0-back_ = 0.331, *SE* = 0.056, *t*(129) = 5.85, *p* < .001, *ß*_2-back – 1-back_ = 0.633, *SE* = 0.056, t(129) = 11.19, *p* < .001) as post-hoc tests revealed.

**Figure 2.**
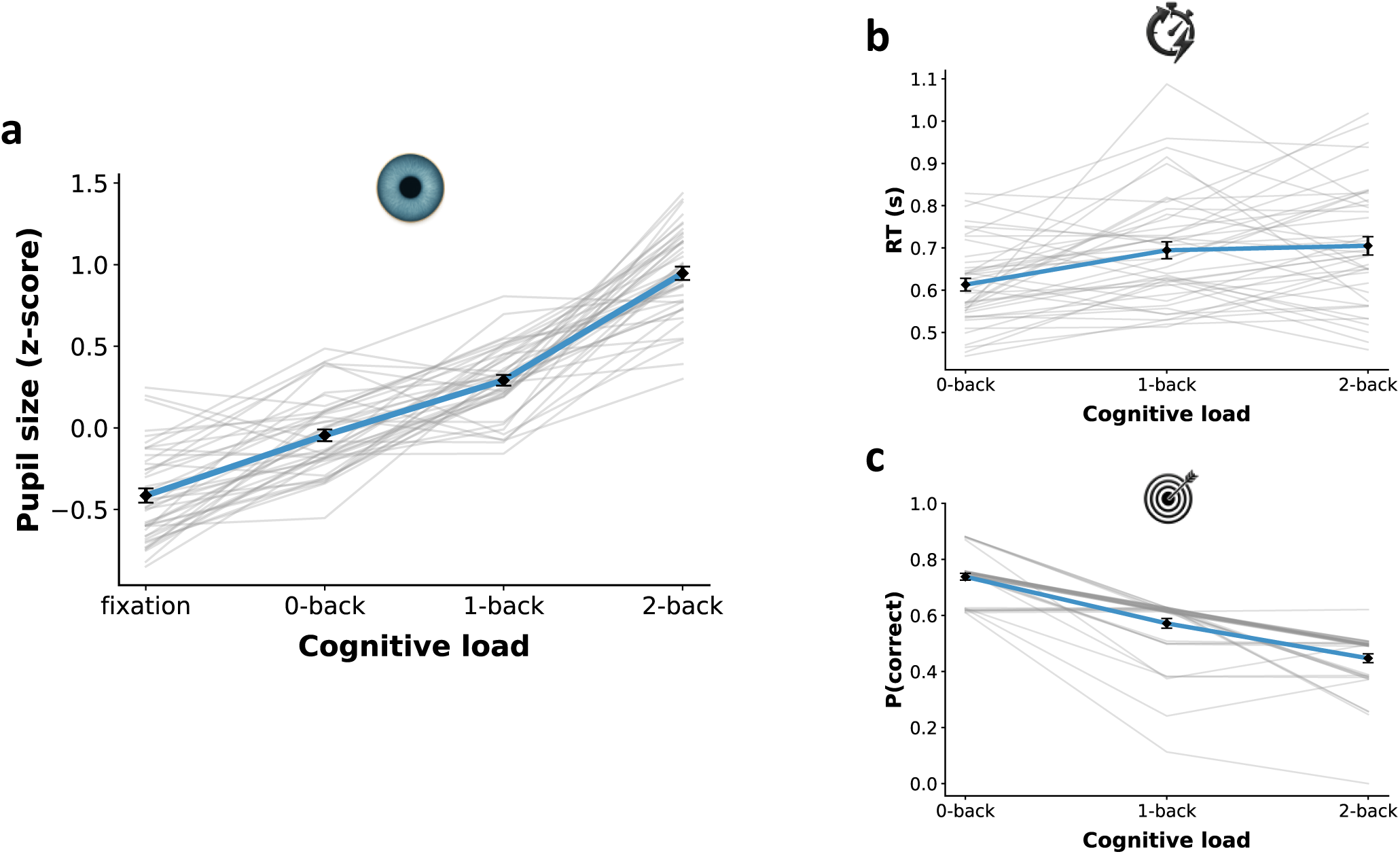
Pupil size, reaction time and accuracy data shown as a function of varying cognitive load levels: fixation, 0-back, 1-back and 2-back, where fixation did not include a behavioral response. Grey lines represent the individual participant data, where the blue line shows the mean course over the different cognitive load levels. Diamonds represent the mean for each cognitive load condition and black error bars reflect the standard error of the mean. This is shown for **a** mean pupil size values over the respective six second time window for each cognitive load condition **b** mean reaction time values for the correctly answered targets and **c** mean accuracy data, defined as the proportions of the number of correct responses out of the eight targets per cognitive load condition block. Note that for the accuracy data, uniformly distributed noise in a range of ± 0.01 was added to the individual data (grey lines) to provoke a slight shift for visualization purposes only (thicker grey line) as the majority of participants had identical values, obscuring individual line traces.

For the behavioral data, we fitted a linear mixed effect model for reaction time (**Figure 2b**). Reaction times increased significantly as a function of cognitive load (*ß* = 0.047, *SE* = 0.01, *F*(1, 86) = 23.64, *p* < .001), meaning participants were slower as cognitive demand increased (*M*_0-back_ = 0.613, *SE*_0-back_ = 0.019, *M*_1-back_ = 0.692, *SE*_1-back_ = 0.019, *M*_2-back_ = 0.707, *SE*_2-back_ = 0.019). Pair-wise post hoc tests showed that only the difference between the 1-back and 0-back condition were significant (*ß*_1-back – 0-back_ = 0.079, *SE* = 0.019, t(85) = 4.16, *p* < .001), whereas the difference between 2-back and 1-back was not (*ß*_2-back – 1-back_ = 0.015, *SE* = 0.019, *t*(85) = 0.79, *p* = 0.710). For the accuracy data, we fitted a generalized mixed effect model (**Figure 2c**), indicating a decrease in performance with cognitive load level. Pairwise comparisons showed that all conditions differed significantly from each other indicated by their odds ratio (*OR*_1-back – 0-back_ = 2.08, *SE* = 0.34, z = 4.46, *p* < .001, *OR*_2-back – 1-back_ = 1.67, *SE* = 0.26, z = 3.35, *p* = .002). Crucially, the differential effects of increasing cognitive load were not as pronounced for the behavioral measures (**Figure 2b and c**) as for our proposed digital biomarker – mobile pupillometry in experimental conditions (**Figure 2a**) – underscoring its high sensitivity.

### Test-retest reliability of cognition-assessed mobile pupillometry

To test the reliability and hence stability of cognition-assessed mobile pupillometry over time (two to three months), participants performed the experimental procedure a second time under identical conditions (same set-up and time as on the first measurement). We computed three stability metrics between day one and day two measurements, each adding a different layer of information for the stability of our measure, commonly required for biomarker validation (**Figure 3a and b**). First, the Pearson correlation indicated a strong, positive and significant association (*r*(31) = 0.74, *p* < .001, 95% CI [0.53, 0.86]), demonstrating temporal stability in the magnitude of pupil response across participants. Second, to test the stability of pupil size rankings across measurements, we determined the Spearman rank coefficient, showing a strong, positive and significant monotonic relationship between participants day one and day two pupil response profile (*r_s_*(31) = 0.80, *p* < .001). In other words, this high degree of rank stability indicates that most participants remained at their position relative to each other, most participants with lower pupil responses in the first assessment also had a lower response in their re-assessment approximately 60 to 90 days later, while the same holds for those with stronger pupil responses. Third, we assessed the test-retest reliability using a two-way mixed-effects single-measures ICC showing good (Cicchetti, 1994; Koo & Li, 2016) reliability (ICC = 0.73, 95% CI [0.53, 0.86]). These three metrics provide strong evidence for good stability over several months of mobile-assessed pupil response profiles as participants’ response magnitudes were consistent, their relative ranking was preserved and the absolute measurements were reliably reproducible across sessions.

**Figure 3.**
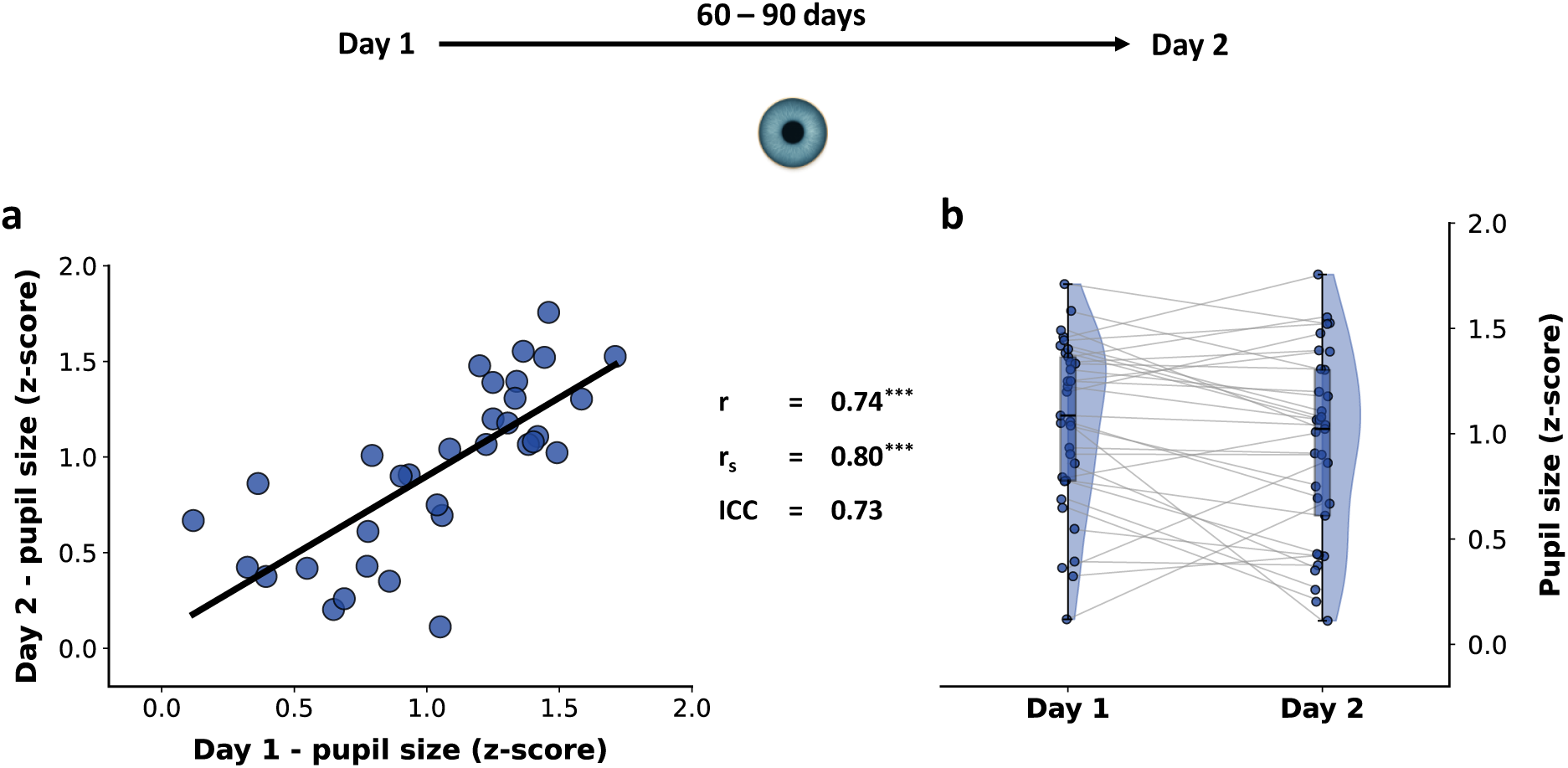
Test-retest reliability of differential pupil size score of 2-back and 0-back. Data is shown for all participants that were measured twice with 60 to 90 days apart between measurements. This is shown as **a** correlation between day 1 and day 2 and **b** boxplots with individual pupil values for day 1 and day 2 connected by grey lines including overall data distributions for each day. We report the Pearson correlation coefficient (r), Spearman rank correlation coefficient (r_s_) and Intraclass Correlation Coefficient (ICC).

### Clinical validity of cognition-assessed mobile pupillometry

In order to examine the clinical validity of mobile pupillometry with VR as digital biomarker, we tested three different clinical cohorts with varying levels of cognitive impairment and compared them to healthy control participants. In **Table 1**, the clinical and demographic characteristics of each cohort are shown.

As shown in **Figure 4a**, there was a significant main effect of cognitive load condition (*F*(3, 498) = 289.42, *p* < .001). There was also a significant main effect of group (*F*(3, 166) = 9.50, *p* < .001). Importantly, there was a significant interaction between condition and group (*F*(9, 498) = 10.50, *p* < .001), indicating that the effect of condition differed across groups. Given the significant interaction, we performed post-hoc comparison, to unravel the group differences across task conditions. During fixation, patient’s pupil response profile of the cognitively impaired groups, i.e., ODI-CC (*estimate* = −0.316, *SE* = 0.067, *p* < .0001) and Memory Clinic (*estimate* = −0.220, *SE* = 0.084, *p* = .045) significantly differed from HC. During low cognitive load (0-back), differences between groups were small, only the BeCOME group had significantly lower pupil size than HC (*estimate* = 0.161, *SE* = 0.050, *p* = .007). With increasing load as in the 1-back, the cognitively impaired groups (ODI-CC and Memory Clinic) showed an increased pupil size which differed significantly to the BeCOME group (*estimate* = −0.253, *SE* = 0.070, *p* = .002 and *estimate* = −0.288, *SE* = 0.087, *p* = .006). The pupil size of the BeCOME group was again also significantly lower than of HC (*estimate* = 0.177, *SE* = 0.050, *p* = .002). During highest cognitive load, group differences were most pronounced. The BeCOME group had a similar pupil size as HC, which was still significantly lower (*estimate* = 0.130, *SE* = 0.050, *p* = .044). The strongest cognitively impaired patient groups (ODI-CC and Memory Clinic) showed the smallest pupil size, which were significantly lower compared to HC (*estimate* = 0.420, *SE* = 0.067, *p* < .0001 and *estimate* = 0.410, *SE* = 0.084, *p* < .0001). Both groups, ODI-CC and Memory Clinic did not differ from each other (*p* = .999).

**Figure 4.**
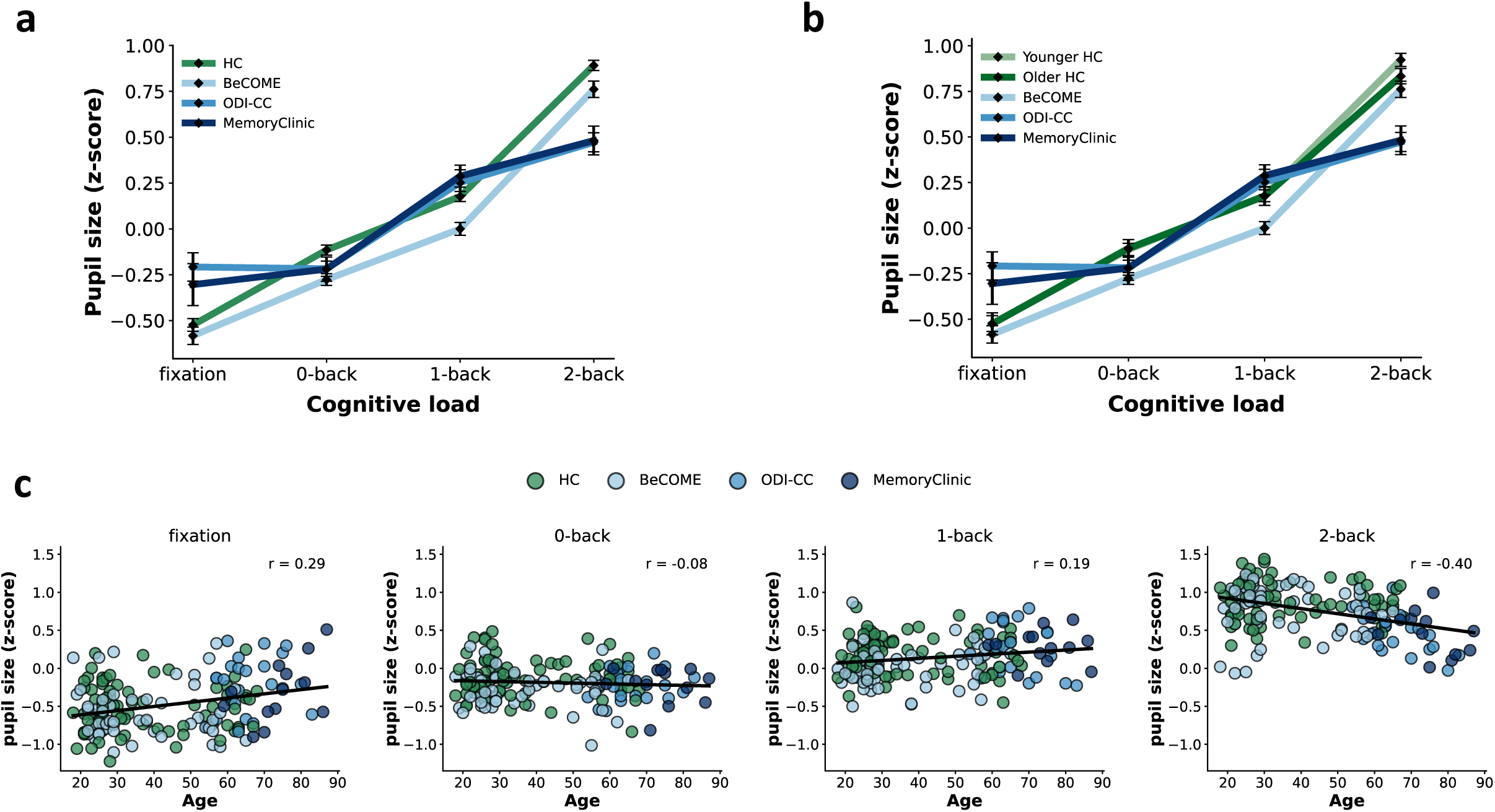
Pupil response profile across patient cohorts and its relation to age. The individual lines show the mean course over the different cognitive load levels. Diamonds represent the mean for each cognitive load condition and black error bars reflect the standard error of the mean. **a** Pupil responses for each patient cohort and HC as a function of cognitive load **b** Pupil responses for each patient cohort as a function of cognitive load with HC split in younger HC (below 40 years of age) and older HC (40 years or older) group **c** Correlations of pupil dilation and age for each cognitive load level.

Given that we were interested in the influence of age on cognitive impairment level and to which extent this relates to our pupil marker (**Figure 4b, c**), we fitted another model with the same specifications as before but adding age as term and coding condition as as continuous predictor with its ordered levels representing increasing cognitive load. The model revealed a signifanct three-way interaction between age, cognitive impairment group and condition (*F*(3, 502.00) = 3.57, *p* = .014), indicating that the effect of condition on pupil size differed across groups and varied as a function of age. In addition, there was a significant interaction between condition and age (*F*(1, 502.00) = 13.37, *p* < .001), indicating that the influence of condition on pupil responses changed with age. Furthermore, a significant group × age interaction was observed (*F*(3, 643.16) = 2.95, *p* = .032), suggesting differential age effects between groups. There were significant main effects of condition (*F*(1, 502.00) = 139.14, *p* < .001) and age (*F*(1, 643.16) = 10.72, *p* = .001), indicating overall differences in pupil size across conditions and age. Note that the elderly healthy participants scored very similar to the younger healthy participants, and dissimilar to the elderly patients (**Figure 4b**), indicating that the results are not due to various age groups across the cohorts.

## Discussion

In the present study, the main goal was to validate mobile pupillometry with VR under experimental conditions as novel digital biomarker for cognitive function and impairment. For this, we assessed the pupil signal during working memory processing, operationalized with the n-back task in a VR environment. We investigated specifically three main required properties for such a potential mobile biomarker, namely its construct validity, its reliability and its clinical validity.

First, to test its construct validity, our results confirmed that pupillometry in VR provides sensitive differences in pupil size as a function of cognitive load level, that is, pupil size increased with increasing working memory demands. These differences were as strongly pronounced as with commonly lab-based high-resolution eye-tracker systems (Fietz et al., 2024; Yeung et al., 2021). In contrast, for the behavioral metrics (i.e., RT and accuracies), the differences across cognitive load levels were far less pronounced with much more variability obscuring sensible differential responses to different working memory load. These results point towards a higher sensitivity of mobile pupillometry in capturing meaningful cognitive differences than behavioral readouts alone.

Second, to assess its reliability, participants performed the same n-back working memory task 60 to 90 days after the initial measurement. We observed a highly stable pupil response pattern in participants both analyzed in terms of their rank position within a group and in terms of their absolute pupil magnitude. In order to derive meaningful information from a biomarker, it has to be reliable, otherwise no consistent conclusions and hence no useful information can be drawn. In this context, our reliability analyses provided converging evidence that mobile pupillometry is a reliable digital biomarker for assessing the cognitive domain of working memory function.

Third, with respect to its clinical validity, we observed that mobile pupillometry assessment was sensitive enough to reveal differential profile patterns between patient cohorts with varying severely affected cognitive impairment and healthy control groups. The cohorts with the strongest cognitive impairment level (i.e., ODI-CC and Memory Clinic) showed the weakest increase in pupil size and, therefore, potentially the weakest LC-NA upregulation when cognitive effort was highest (as in the 2-back condition).

Noteworthy, both cognitively impaired patient cohorts exhibited the largest pupil size in the 1-back condition compared to the other groups, likely reflecting an overshooting response in which the affected LC-NA system allocates a maximal arousal upregulation response, requiring substantial physiological resources. This finding is in line with literature demonstrating compensatory, overshooting responses in physiological systems in patients with MCI and AD (David et al., 2025; Granholm et al., 2017). However, this highly unsustainable state appears to fail when physiological demand due to increased cognitive effort becomes too high, as in the 2-back condition, which may explain the dampened response observed in our data.

As the LC is among the first brain regions affected by tau pathology in early stages of AD (Gutiérrez et al., 2022), it is a possibility that patients in our ODI-CC and Memory Clinic groups exhibited dysfunction in this system. Interestingly, the pupil size profile for the high-functioning BeCOME group resembled the profile of the healthy control group with overall attenuated magnitudes across cognitive load conditions. This may indicate a physiological system that still functions appropriately in response to increasing cognitive demand, yet with potentially reduced arousal regulation capacity (Benning & Ait Oumeziane, 2017; Brendler et al., 2024; Fietz et al., 2024). However, overall reduced values in the BeCOME group could also be at least partially explainable by minor experimental procedural differences, given that these patients underwent a 6-minute fixation before the n-back task and completed eye calibration prior to the first task.

The fact that mobile pupillometry revealed variable response profiles as a function of cognitive impairment level underscores its sensitivity and its potential to capture the continuum from healthy cognitive status to early neurodegenerative disease. Another key finding was that reduced pupil size as a function of cognitive load and cognitive impairment was dependent on age. This suggests that normative decline in the LC-NA arousal system due to physiological aging may exacerbate the effects of clinical cognitive impairment. It may be speculated that increasing age worsens cognitive impairment because compensatory mechanisms can no longer be adequately recruited. These findings highlight the importance of early prevention of cognitive impairment, as older age appears to further compromise the physiological system, particularly in affected individuals, indicating a need for age-specific normative pupil values for comparison. We also observed increased pupil size during fixation, which was stronger in cognitively impaired patients than in HC. As no information had to be actively maintained, this finding of increased allocation of the LC-NA system cannot be attributed to working memory respectively cognitive load. Rather, cognitive impairment could have been a moderator for different attentional and fatigue levels in patients compared to healthy controls, which are processes that are active during fixation (Zhang et al., 2023). Together, these results indicate that mobile pupillometry offers an assessment that taps into the dysfunctional neurophysiological system in patients, who fail to provide the required arousal level for required cognitive demand, thereby supporting its utility as a sensitive measurement tool for early cognitive impairment.

Integrating the previously discussed findings and embedding them into the digital biomarker literature (Coravos et al., 2019; Powell, 2024; Vasudevan et al., 2022; Wright et al., 2017), we demonstrated that mobile pupillometry may constitute a valid and reliable digital biomarker. There are certain additional features being discussed in the field and assigned high importance. A recurrent topic in biomarker discussions is its safety for the individual (Coravos et al., 2019). Our measure is completely non-invasive and poses no safety concerns for the tested individual. However, one has to acknowledge that it is not suited for people with eye or neurological conditions (e.g., epilepsy, migraine, etc.) as the VR scenarios could provoke undesirable reactions in these conditions. Nonetheless, with respect to its application value, our results indicate that mobile pupillometry during cognitive tasks holds promise as digital biomarker for cognitive functioning. As we have shown its construct validity, it could be used as working memory screening tool offering meaningful biological information concerning the functional state of the underlying LC-NA arousal system. Furthermore, as we demonstrated its high reliability over several months, it provides ideal conditions to be used as a monitoring tool for cognitive status and impairment. Noteworthy, as disease-modifying pharmacological interventions for neurodegeneration such as AD become more important (e.g., Lecanemab; Van Dyck et al., 2023), it could aid biologically-informed decisions on therapeutic protocols or serve as monitoring marker. Overall, we provided initial evidence in this validation that VR pupillometry under experimental conditions constitutes a reliable and valid digital biomarker for assessing cognitive functioning and impairment across the spectrum from health to disease. Furthermore, we provide the basis for this mobile pupillometry set-up to be used in other experimental contexts and study populations.

Nonetheless, our promising findings should be viewed within the framework of the study limitations. Even though we observed strong effect sizes, biomarker validation requires larger sample sizes. This holds specifically for the clinical validation sample. Future research should build on our initial validation results in a larger cohort to underline the robustness of these findings. Furthermore, one has to acknowledge that we studied cognitive impairment from a relatively homogenous perspective by specifically focusing on the working memory component, which we were able to physiologically capture with mobile pupillometry. Cognition and therefore cognitive impairment, however, encompass additional processes such as attention, executive and visuospatial function, yielding a complex mix of deteriorated processes together with working memory difficulties. In the present study, we did not test particularly for these various combinations and thus our results should only be interpreted within the working memory context. Future research could address this point by expanding on meaningful syndrome profiles by clustering patients based on similarities in cognitive deficits, allowing to draw meaningful conclusions of this digital marker and how physiological processing is affected. This could eventually lead to a more personalized approach and informed clinical decisions. Furthermore, we have shown that assessments of cognitive functioning with mobile pupillometry remains highly stable over several months, qualifying for monitoring of a decline in cognition. However, in contrast to the HC cohort, we measured patients only on a single occasion and thus the extent to which the pupil response profile changes in the light of progressive memory deterioration or worsening of clinical course can only be speculated on. Therefore, future studies should implement a longitudinal mobile pupillometry follow-up design to systematically monitor patients over time.

In conclusion, in this technical and clinical validation of mobile pupillometry under experimental conditions, we provide initial evidence for mobile pupillometry being a valid and reliable digital biomarker for assessing cognitive functioning in healthy, neuropsychiatric and neurodegenerative cohorts. Our results demonstrate excellent characteristics to capture arousal-related working memory processes that remained stable over several months in healthy participants. Furthermore, mobile pupillometry is sensitive to disruptions of working memory processes in affective and neurodegenerative disease and provides neurobiological information beyond behavioral measurements. We envision that mobile pupillometry with its location-independent set-up and excellent digital biomarker characteristics finds its use in assessing individuals’ cognition status and those at risk of, or suffering from, cognitive impairment, thereby efficiently guiding clinical decisions and relieving a strained healthcare system.

## Data Availability

The data of the current study are not publicly available given the sensitive nature of patient data and associated privacy concerns but can be made available in an anonymized form from the corresponding author on reasonable request and approval by appropriate institutional ethics committee.

## Acknowledgements

The authors would like to thank the Psychiatric Study Center of the Max Planck Institute of Psychiatry - Dr. Norma Grandi, Anna Gossmann, Elisabeth Kappelmann, Karin Hofer, Alexandra Kocsis, Vlada Kolysnik and Gertrud Ernst-Jansen - for organizational support, biomaterial and data collection and the Biomaterial Processing and Repository Unit of the Max Planck Institute of Psychiatry - Tamara Namendorf, Božidar Novak, Marketa Reimann, Angelika Sangl - for the processing and storage of the study’s biosamples. The authors further thank Alexandra Bayer, Ines Eidner, Anna Hetzel, Elke Frank-Havemann, Viktoria Messerschmidt, and Ursula Ritter-Bohnensack for assisting with MRI scanning and Julia- Carolin Albrecht, Anastasia Bauer, Anja Betz, Alina Feichtinger, Rebekka Hirsch, Miriam El- Mahdi, Eila Mertens, Carolin Haas, Katharina Kahn, Lisa Kammholz, Sophia Koch, Christof Leininger, Anna Lorenz, Rebecca Meissner, Jessie Osterhaus, Liisbeth Pirn, Christina Rieger, Linda Schuster and Refican Yasar for their help with data collection, study management, the recruitment, and screening of BeCOME participants. The authors’ special thanks go to all study participants for participation in the BeCOME study.

## BeCOME Working Group

Elisabeth Binder, Angelika Erhardt, Susanne Lucae, Philipp G. Sämann, Norma C. Grandi, Tamara Namendorf, Michael Czisch, Immanuel Elbau, Laura Leuchs, Anna Katharine Brem, Leonhard Schilbach, Sanja Ilić-Ćoćić, Juliuis Ziebula, Iven-Alex von Mücke-Heim, Yeho Kim, Julius Pape, Michael Gottschalk, Alexandros Balaskas

## Memory Clinic Working Group

Peter Falkai, Sandra Nischwitz, Philipp G. Sämann, Manfred Uhr, Laura Perna, Mirko Endres, Martin Beinsteiner, Laura Fischer, Michael Gottschalk, Katja Bochmann

The presentation of the experimental scenarios such as the n-back task in VR was developed in Unity by Florian Binder under the supervision of Victor Spoormaker, in an extension of previously used VR scenarios (see Binder & Spoormaker 2020; Binder et al. 2022): https://github.molgen.mpg.de/spoormaker/VR-Experimental-Presentation

## Author contributions (CRediT’s)

**Andy Brendler (AB):** Writing – original draft, Conceptualization, Project Administration, Investigation, Methodology, Formal Analysis, Data Curation, Visualization, Software, Writing – Review & Editing; **Julia Fietz (JF):** Investigation, Data Curation; **Anastasia Bauer (ASB):** Investigation, Data Curation, Formal Analysis; **Daniel Pfahl (DF):** Investigation, Data Curation, Formal Analysis; **Sarah Higgins (SH):** Investigation, Data Curation; **Eva Vidovic (EV)**, **Kerstin Hupe (KH)**, **Matthias Knop (MK):** Data Curation; **Tanja Brückl (TB)**: Data Curation, Writing – Review & Editing**; Victor I. Spoormaker (VIS):** Conceptualization, Methodology, Supervision, Project Administration, Writing – Review & Editing, Funding Acquisition. All authors have read and approved the final manuscript.

